# Two Decades of Endemic Dengue in Bangladesh (2000-2022): Trends, Seasonality, and impact of Temperature and Rainfall Patterns on transmission dynamics

**DOI:** 10.1101/2023.07.16.23292380

**Authors:** Mohammad Nayeem Hasan, Ibrahim Khalil, Muhammad Abdul Baker Chowdhury, Mahbubur Rahman, Md Asaduzzaman, Masum Billah, Laila Arjuman Banu, Mahbub-ul Alam, Atik Ahsan, Tieble Traore, Md. Jamal Uddin, Roberto Galizi, Ilaria Russo, Alimuddin Zumla, Najmul Haider

**Author notes:** **Corresponding author (NH):** Dr Najmul Haider, School of Life Sciences, Keele University, Huxley Building, Room 122, Keele, Staffordshire, ST5 5BG, United Kingdom.

## Abstract

**Background:** The objectives of this study were to compare the dengue virus (DENV) infection, deaths, case-fatality ratio, as well as meteorological parameters between the first and and the recent decade (2000-2010 vs. 2011-2022) and to understand the trends, seasonality, and impact of change of temperature and rainfall pattern on transmission dynamics of Dengue in Bangladesh

**Methods:** For the period 2000-2022, dengue cases and death data from Bangladesh’s Ministry of Health and Family Welfare’s website, and meteorological data from the Bangladesh Meteorological Department were analyzed. Mann-Kendall and Sen’s slop tests were used for trends and variations and fitted a time series Poisson regression model to identify the impact of meteorological parameters on the incidence of dengue cases. A forecast of dengue cases was performed using an autoregressive integrated moving average model.

**Results:** Over the past 22 years, a total of 244,246 dengue cases were reported including 849 deaths (Case fatality ratio [CFR] =0.34%). The mean annual number of dengue cases increased eight-fold during the second decade, with 2216 cases during 2000-2011 vs. 18,321 during 2012-2022. The mean annual deaths have doubled (21 vs. 46) although the overall CFR had decreased to one-third (0.69 vs 0.24). Between the periods, the annual temperature increased by 0.49 °C, and rainfall decreased by 314 mm despite increasing unusual rainfall in the pre-and-post monsoon period. An increasing trend of dengue cases is observed with a much stiffer rise after 2018. Monthly mean temperature (Incidence risk ratio [IRR]: 1.26), first-lagged rainfall (IRR: 1.08), and second-lagged rainfall (IRR: 1.17) were significantly associated with monthly dengue incidence.

**Conclusions:** The increased local temperature and unusual rainfall might have contributed to the increased incidence of DENV infection in Bangladesh. Community engagement, vector control, and destruction of mosquito habitats are key to controlling dengue.

## Introduction

Dengue fever is a mosquito-borne diseases (MVD) caused by four distinct serotypes of the dengue virus (DENV) of the Flaviviridae family ^1^. DENV is transmitted to humans by the bites of the female Aedes species mosquitoes including *Ae. aegypti* and *Ae. albopictus* ^1,2^. DENV is endemic in over 125 countries of the world and the number of cases globally reported to WHO continues to increase every year ^3^. Annually, an estimated 390 million dengue infections are recorded across the world, including 96 million clinical cases ^4,5^. Most infections (>80%) are self-limiting with no or mild clinical manifestation resulting in lifelong immunity for serotype ^6^. However, infections with different serotypes, known as secondary dengue infection, may result in severe dengue with a higher case-fatality ratio^7^.

Currently, South and Southeast Asia is considered to be the hotspot of DENV infection with more than 50% of cases recorded in the regions ^8^. The first official DENV outbreak in Bangladesh was reported in 2000, and since then, dengue has become endemic in the country posing a significant health challenge ^9^. Over the past few years, the number of dengue cases has been steadily increasing with significant seasonal and regional variations. Analysis of data from 2000 to 2017 revealed that almost half of the dengue cases occurred during the monsoon season (May-August) and the post-monsoon season (September-December) ^10^. However, a shift in seasonal patterns has been observed since 2014, with dengue cases being reported during the pre-monsoon season as well ^10^. During 2015-2017, the number of dengue cases during the pre-monsoon season was more than seven times higher compared to the previous 14 years ^10^.

Climate change including changes in precipitation, temperature, and humidity, as well as rapid unplanned urbanization, were identified as strong predictors of an ecological imbalance that has led to an increase in dengue cases in Bangladesh ^10^. This suggests that the dengue transmission season could eventually extend year-round, with a higher chance of outbreaks occurring at any time of the year. Identifying trends and seasonality in dengue cases can aid health authorities and relevant public and private administrations in effectively allocating resources to control the spread of the DENV through vector control. The objectives of our study were to: i) compare the annual and monthly cases in the first [2000-2010] and recent decade [2011-2022], ii) identify the trend and seasonality of dengue cases, iii) quantify the impact of climatic parameters for the monthly incidence of dengue cases in the country and iv) forecast the annual incidence of dengue cases for next decade.

## Methods

### Data sources

The data on the number of reported dengue-infected people have been extracted from the Directorate General of Health Services (DGHS)’s website from January 2000 to December 2022^11^. We used the definition of dengue cases used by the Ministry of Health and Family Welfare, Bangladesh, which was discussed in our earlier article ^12^. We collected three-hourly temperature and daily rainfall data from Bangladesh Meteorological Department (BMD) over the period 2000–2022 ^13^ for the meteorological station located in Mirpur, Dhaka.

### Variables

The monthly number of dengue cases was used as the primary outcome variable. Two climatic variables-temperature and rainfall are used as the covariates for the regression analysis. In addition, two lagged variables rainfall in lag 1 and lag 2 have also been used as the predictors for the incidence of monthly dengue cases to capture the actual impact of those meteorological parameters. We also used monthly mortality data for comparison between two decades.

### Statistical analysis

We analyzed the monthly dengue incidence and meteorological data for the period of 2000-2022. In the first stage, descriptive analysis was conducted to determine the characteristics of confirmed dengue cases and deaths with mean, and standard deviation in each year and each month for the entire period. Then, we compared dengue cases, deaths, and weather parameters in two decades (2000-2010 and 2011-2022) using paired sample t-test. Next, we calculated the monthly growth factor (GF) of dengue cases by dividing the number of dengue cases reported in each month by the number of dengue cases reported in the previous month and repeating this process for each month from 2000 to 2022 ^14^. The formula for the growth factor can be given by

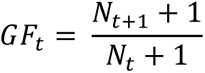

where *N_t_* indicates the number of dengue cases in *t*th month. To avoid the occurrence of zeros in some months, we added 1 to the total number of cases for each month. This allows us to obtain a real-valued measurement of the GF for the above equation. The distribution of GF was skewed; therefore, we used the first natural log transformation before the data was further examined.

However, we have also performed a reverse transformation of the log (GF) values by exponentiating values to convert them to the original scale for ease of interpretation^14^.

We performed forecasting using the autoregressive integrated moving average (ARIMA) model. The ARIMA model is a data-driven, exploratory strategy that enables us to fit a suitable model and forecast values. The ARIMA model consists of autoregressive (p) terms, differencing (d) terms, and moving average (q) operations, and it is denoted as ARIMA (p, d, q) ^15^. To select the appropriate autoregressive and moving average orders, the autocorrelation function (ACF) and partial autocorrelation function (PACF) were examined. Additionally, the differencing parameter, represented by “d,” indicates the number of times the time series is different to achieve stationarity. An ARIMA (p, d, q) process refers to an autoregressive moving average (ARMA) model that has been differenced “d” times to obtain stationarity (Hasan et al. 2021). By removing high-frequency noise from the data, the model discovers local patterns by assuming that the time series values are linearly related. We also conducted a Mann-Kendall (M-K) trend analysis to determine possible upward or downward trends ^17^. We also performed the Sen’s slope test to assess variations in annual dengue cases and deaths^18^.

We, then used a time series count generalized linear model (GLM), more specifically, a time-series Poisson regression model to determine whether the climatic factors were associated with the dengue cases over time ^19^. The non-normality, heteroscedasticity, and non-linearity that characterize count data can be fitted easily using GLMs. The time-series observations may possess autocorrelation and they might be nonnegative integers, and thus GLM is useful in overcoming both issues [20, 21, 22]. Monthly dengue cases were utilized as the outcome variable in this model, along with data from the Bangladesh Meteorological Department (BMD) on temperature and rainfall. To capture the actual impact of rainfall on dengue incidence across time, we additionally employed two lagged variables of meteorological elements, mainly rainfall in lag 1 and 2. After eliminating predictors with a higher multicollinear relationship, we have arrived at average temperature, rainfall (in lag 1), and rainfall (in lag 2) as the final set of predictors for the monthly dengue incidence in Bangladesh. We used the statistical program RStudio, version 3.5.2.2 for the analyses ^20^.

## Results

Between 2000 and 2022, Bangladesh reported a total of 244,246 DENV infection with an annual mean of 10,161 cases (± standard deviation [SD]=23,971) including 849 fatal outcomes indicating a case-fatality ratio (CFR) of 0.34%. Between 2000 to 2010, the mean annual number of DENV infection was 2,216 (±2,123) which has increased over eight folds compared to the following decade (2011-2022) at 18,321 (±31,778) **(Table 1)**. Between these two periods, the mean number of annual deaths due to DENV infection has increased by 2.2 times (21.18 cases vs 46.58 cases). However, the CFR of DENV infection has decreased to almost one-third between two decades (0.69 vs 0.24) **(Table 1)**.

**Table 1:** Comparison of dengue cases, deaths, and weather parameters between the first (2000-20210) and the recent decade (2011-2022) in Bangladesh.

|  | First decade (2000-2010) | Recent decade (2011-2022) | p-value |
| --- | --- | --- | --- |
| Mean annual dengue cases (±Standard deviation [SD]) | 2216.64 (±2123.62) | 18321.92 (±31,778.90) | 0.219 |
| Mean annual dengue deaths (±SD) | 21.18 (±30.69) | 46.58 (±90.90) | 0.853 |
| Mean Case-fatality ratio (± SD) | 0.69 (±0.79) | 0.23 (±13) | 0.08 |
| Mean temperature $^{\circ}$C (±SD) | 26.35 (±0.49) | 26.84 (±0.37) | <0.001 |
| Mean annual rainfall (±SD) | 2078.66 (±459.68)<br>[mm] | 1764.50 (±448.32)<br>[mm] | 0.188 |

The highest monthly average number of DENV infections was recorded in August (n=3407 cases) and the lowest was in February (n=7.3 cases) **(Fig 1)**. The highest number of annual DENV infections was reported in 2019 with 101,354 and the highest number of deaths was recorded in 2022 with 281 deaths, which was 35% of total deaths recorded in the past 23 years in Bangladesh **(Fig 1)**. Most of the dengue-related deaths were recorded after 2018, with more than 69% (n=550) deaths recorded during this time **(Fig 1)**.

**Fig 1:**
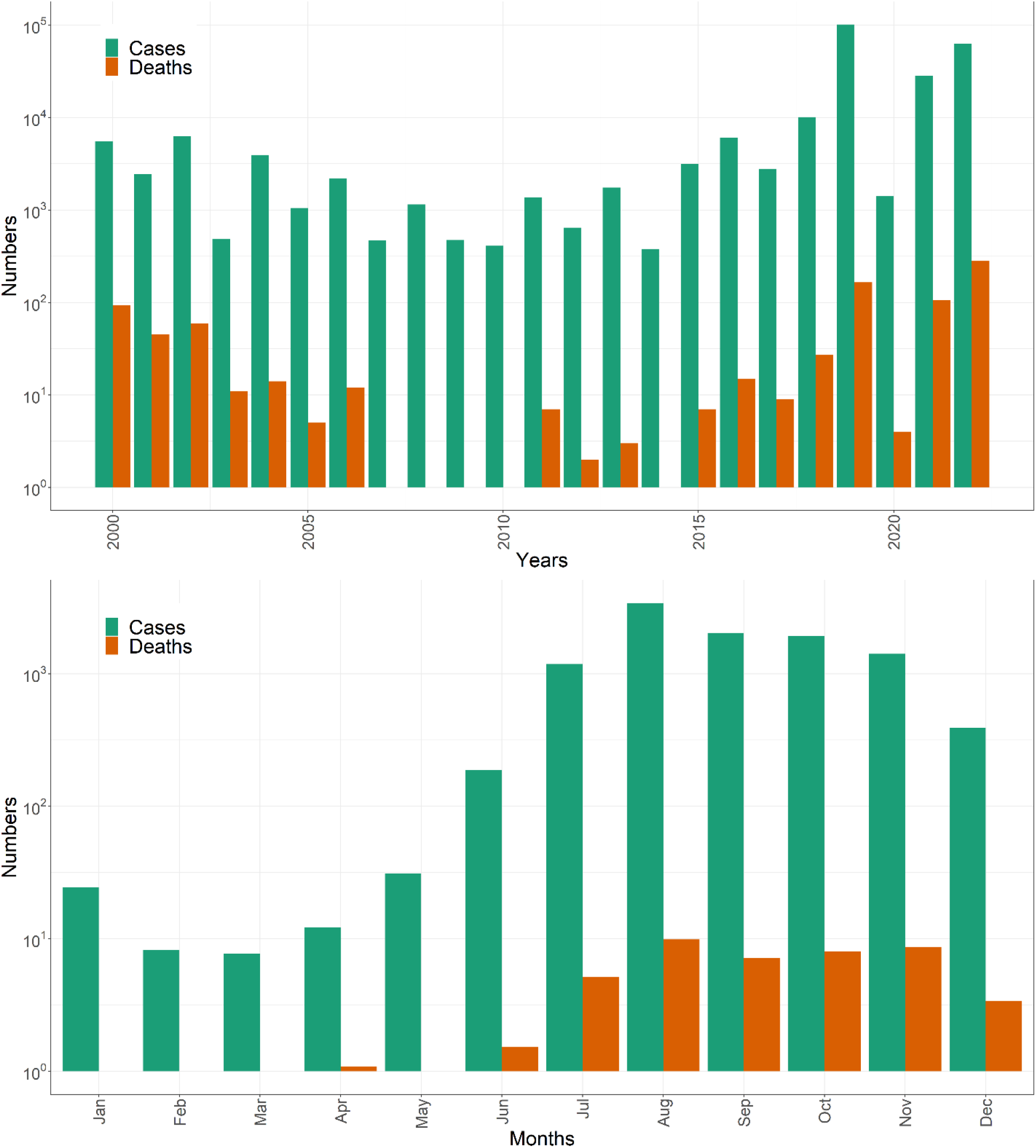
Top: Number of dengue cases and deaths over the period 2000-2022, Bangladesh. Bottom: Number of monthly dengue cases and deaths recorded in Bangladesh.

The average annual temperature was 26.35 °C (SD=3.72) during the first decade (2000-2010) and 26.84 °C (SD=3.76) during the recent decade (2011-2022) (**Table 1**). The increase of 0.49 ° C temperature was equivalent to 4292 degree-hour/year of heat (365 days X 24 hours X 0.49 ° C). The annual rainfall has decreased by 314 mm between two decades (2078.66 mm vs. 1764.50 mm) (**Table 1**), of which 308 mm decreased during the monsoon (July-October) season and only 6 mm decreased during the non-monsoon period. Compared to the first decade (2000-2010), an unusually higher amount of monthly precipitation has been observed in the second decade (2011-2022) with most of the months recording extreme rainfall (more than 3^rd^ quantile value of monthly rainfall for the decade) shown as an outlier of the box plot **(Fig 2)**.

**Fig 2:**
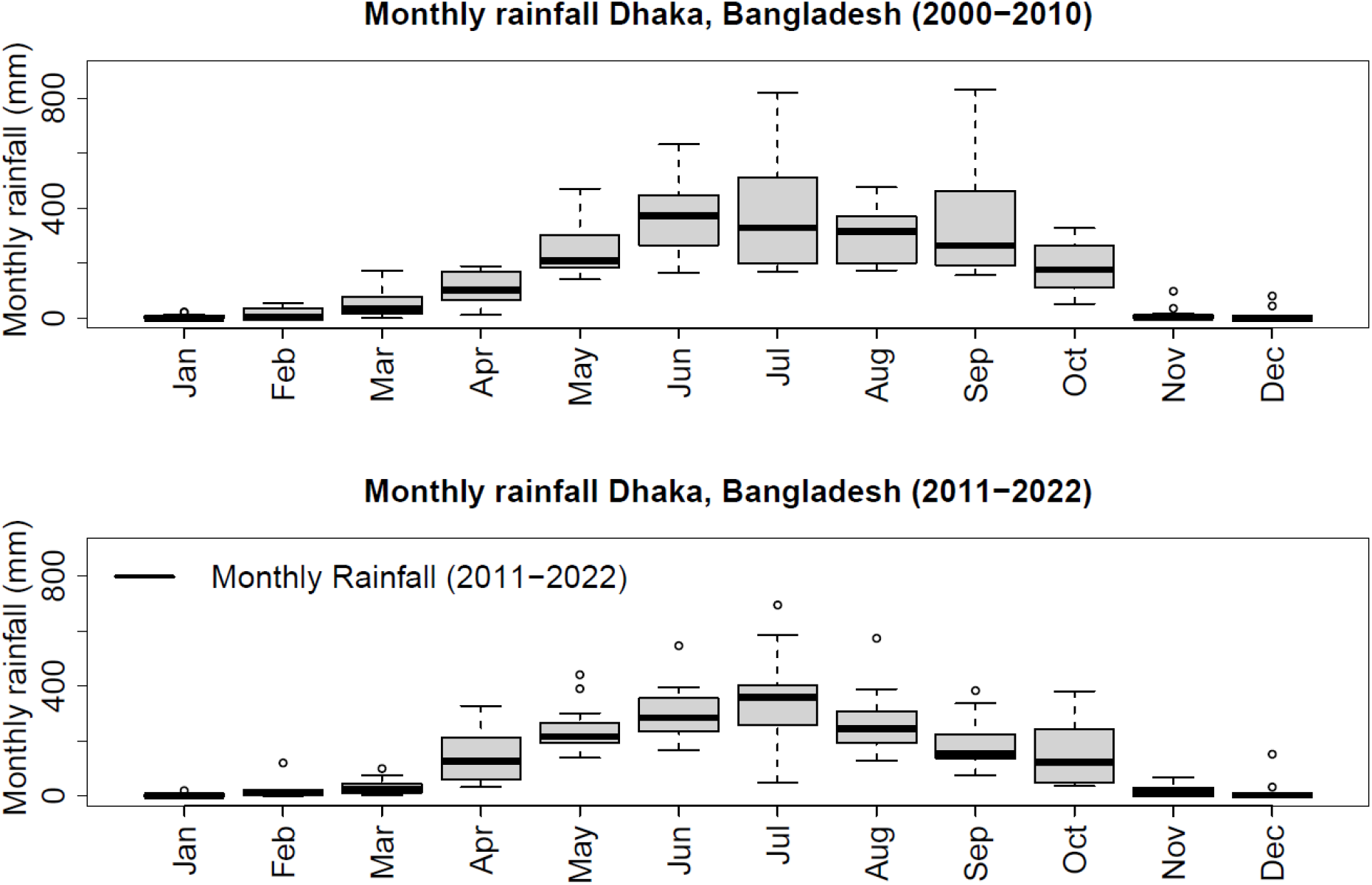
The boxplot compares monthly rainfall in Dhaka city, Bangladesh between two decades. The bottom and top of the box indicate the first and third quantiles, the band inside the box is the median. The dots outside the box are individual outliers. Most of the months in the second decade had outlier rainfall whereas in the first decade, only the cooler months (Nov-Jan) had some extreme rainfall.

The overall mean GF from month to month was 1.37 (SD=0.86). However, in four months (April-July), the monthly GF was above one (lower 95% confidence interval >1), while for the rest of the years, the monthly GF was less than 1 (95% confidence interval crossed 1). More than 77% (71/92) of months between April and July for the period 2000–2022 had mean monthly GF > 1 compared to only 16% (30/184) of months between August and March of the same period. June had the highest GF with a mean value of 3.47 indicating that cases would be more than three times higher in the next month (July). The lowest GF was recorded in December with a mean of 0.54 (95% CI: 0.40 to 0.69) indicating that cases in January would be halved compared to the number of cases recorded in December (**Fig. 3**). In the M-K trend analysis, we found a positive trend of reported dengue cases (p <0.001 and tau = 0.26). In Sen’s slope test, the slope was 171.67 (95% CI: −46 to 687) indicating an upward trend in upcoming months (**Table 2**).

**Fig 3:**
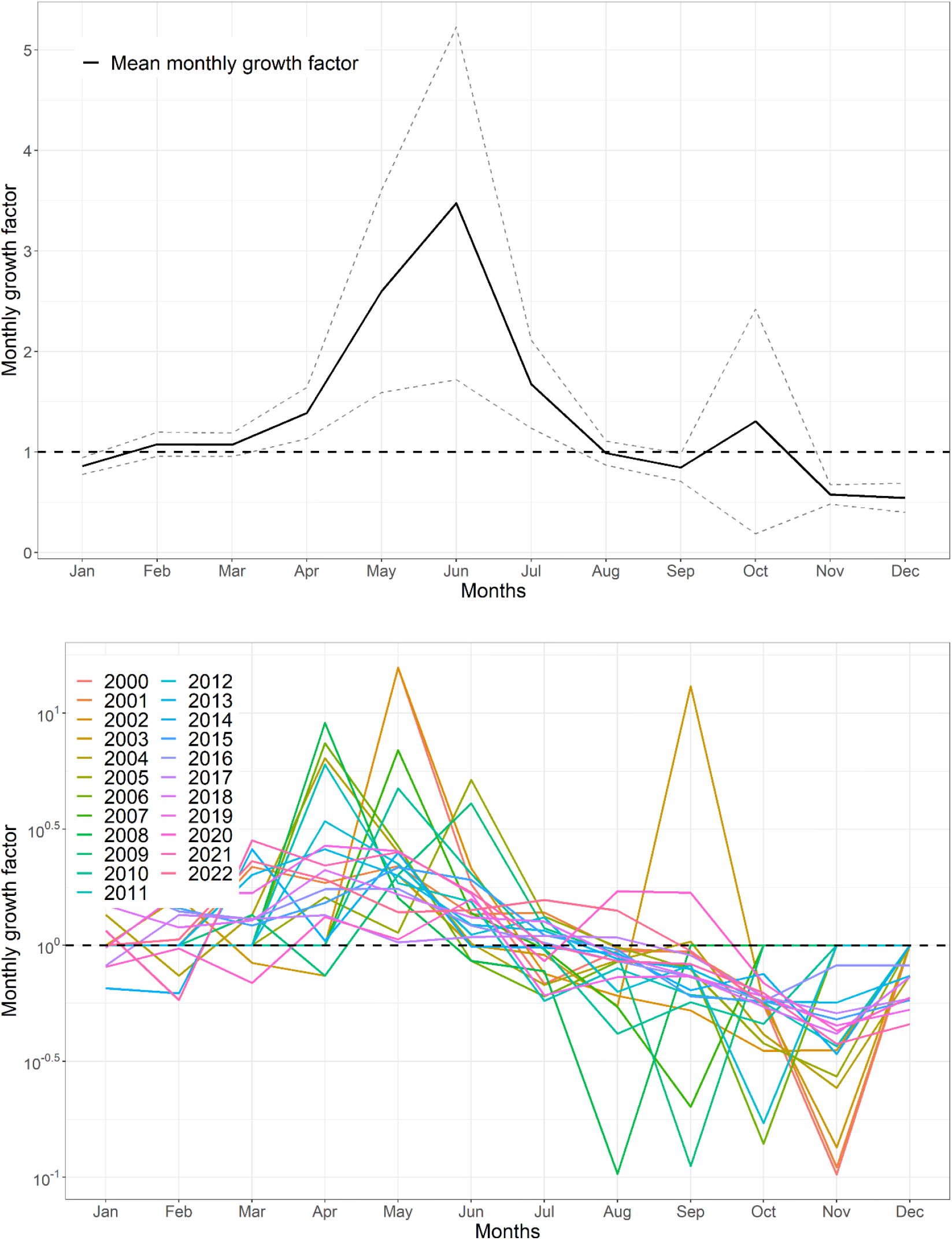
**Top:** Mean monthly growth factor for the period of 2000-2022. Bottom: The Monthly growth factor for the individual year 2000-2022. The dotted horizontal line indicates monthly growth factor 1 (same number of cases in two subsequent months).

**Table 2:** The Mann-Kendell trend test of dengue cases in Bangladesh.

| Test |  |  |
| --- | --- | --- |
| *Mann-Kendell trend analysis* | **Tau** | **p-value** |
|  | 0.26 | 0.139 |
| *Sen's Slop test* |  |  |
|  | Sen's Slope | 95% Confidence Interval |
|  | 171.67 | -46 to 687 |

In the GLM, the estimated effect of each variable is presented as the incidence risk ratio (IRR). The model suggests that dengue cases would rise by 26% for a one-degree centigrade (°C) temperature increase. For each additional centimeter (cm) of rainfall in the first lagged month, the number of dengue cases increased by 8% (IRR= 1.08 [95% CI: 1.07-1.09]), and in the second lagged month increase the cases by 17% [IRR=1. 17 (95% CI: 1. 16 −1.18)] **(Table 3)**.

**Table 3:** The incidence risk ratio (IRR) of average temperature and rainfall to Dengue cases in Bangladesh using time-series count Generalized Linear Model.

|  | IRR (95% CI) | P-value |
| --- | --- | --- |
| Average temperature | 1.26 (1.258 – 1.265) | <0.001 |
| Rainfall (lag 1) in centimeter | 1.08 (1.079 – 1.086) | <0.001 |
| Rainfall (lag 2) in centimeter | 1.17 (1.168 – 1.175)) | <0.001 |

In the ARIMA model, we detected an increasing trend for the first few years, which then started to decline. However, a stiff rise in cases was observed after 2018 except for 2020 (the first year of the Covid-19 pandemic). The forecasted value showed a continuously increasing trend of DENV infection in Bangladesh **(Fig 4)**.

**Fig 4:**
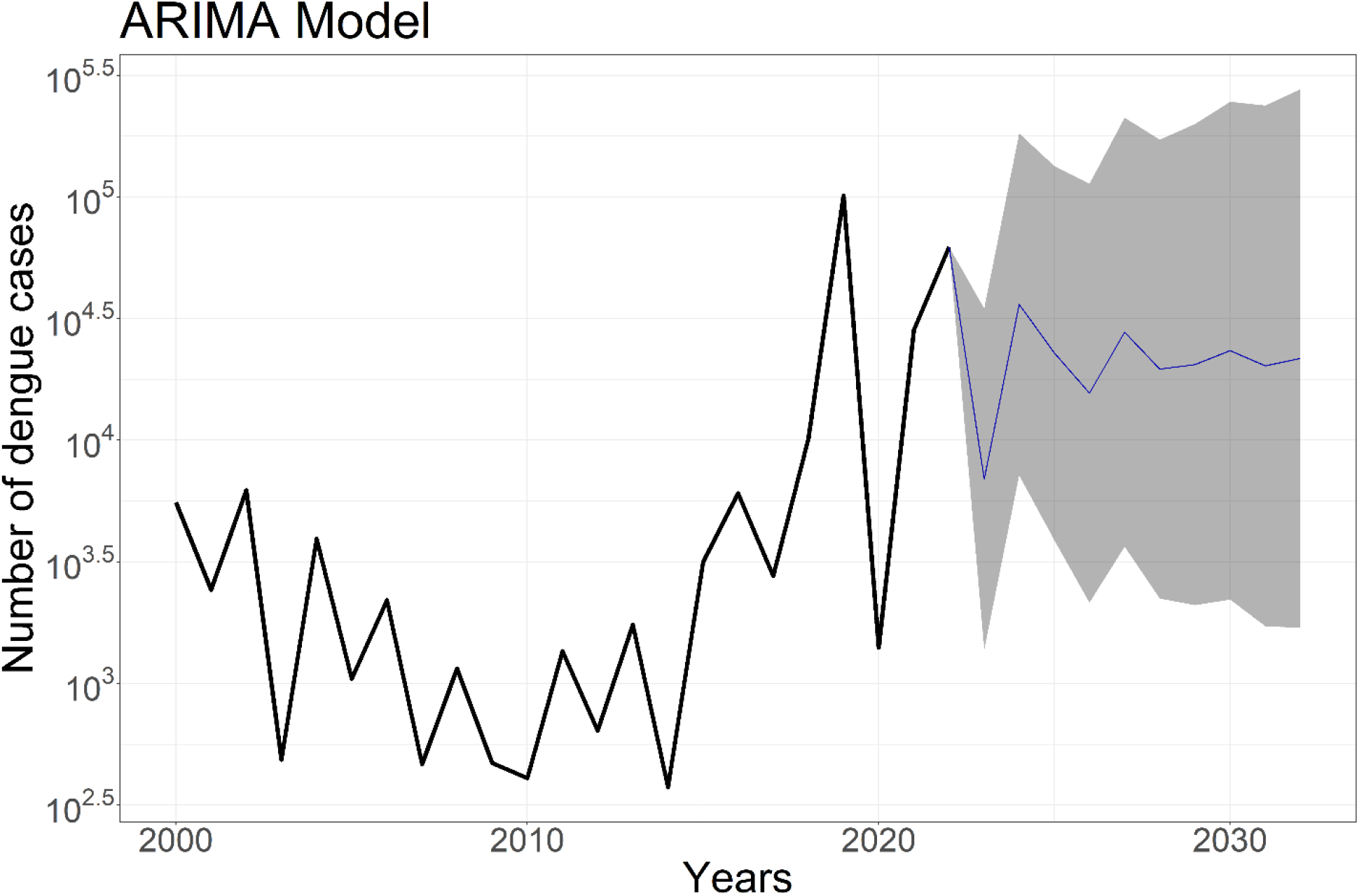
**Fig 3: Top:** Mean monthly growth factor for the period of 2000-2022. Bottom: The Monthly growth factor for the individual year 2000-2022. The dotted horizontal line indicates monthly growth factor 1 (same number of cases in two subsequent months).

## Discussions

Dengue is currently a worrying and important public health challenge for Bangladesh. Our analysis showed that the number of dengue cases has increased eight times and deaths have doubled, and the CFR dropped to one-third between the first and second decade of this century in Bangladesh. Between these periods, the annual temperature increased by 0.49 °C, and annual rainfall decreased by 314 mm, despite changes in rainfall patterns with unusually early or late rainfall outside the typical monsoon season in Bangladesh (July-October) ^21^. The monthly growth factor remains above one significantly for four months (April to July) which overlaps the hot and humid period of the year. Monthly mean temperature, monthly first-lagged rainfall, and second-lagged rainfall played a critical role in monthly dengue incidence in Bangladesh.

The increase of 0.49 °C temperature adds approximately 4292-degree-hours equivalent heat per year in the country. This additional heat would favor mosquito-borne disease transmission. For dengue virus transmission, approximately 305-degree-hours equivalent heat is needed to accomplish the extrinsic incubation period in Aedes mosquitoes at 26° C ^22^. Thus, the additional 0.49°C temperature will add the burden of more than 14 generations of infectious mosquitoes in the environment of Bangladesh. An 8-fold increase in dengue cases is an indication of such changes in temperature in the country. Our model identified a significant role of monthly mean temperature with an additional 1 °C temperature increasing the monthly cases by 26%. Earlier studies showed that for every 1 °C increase in temperature, dengue cases increased by 61% in Australia, 12-22% in Cambodia, 5% in Vietnam, and 2.6% in Mexico ^23^.

Rainfall facilitates mosquito breeding and plays an important role in mosquito-borne disease transmission. Although we found a 15% reduction in annual rainfall in the recent decade from the immediate past decade, we found an increase in unusually high rainfall in pre-and-post monsoon season. Our model showed that both the first and the second lagged month’s rainfall increased monthly cases by 8% and 17%, respectively. These findings are consistent with earlier studies in Bangladesh that showed that peak dengue cases occurred two months after the peak rainfall^24^ or an additional rainy day per month increased dengue cases by 6% in the succeeding month ^25^. Similar findings were reported in Vietnam with the dengue incidence being associated with both first and second-lagged months^26^. In Timor-Leste, a 47% increase in dengue incidence was recorded with an additional 1 mm seasonal rainfall increase ^27^. These findings are biologically plausible as rainfall allows approximately two generations of dengue cases over a month. A generation interval is a time difference between a primary human infection and a second human infection originating from the first human case through two bites of the mosquitoes^28^. To accomplish a generation interval the virus and mosquito undergo several phases including intrinsic incubation period in humans, human-mosquito transmission (first bite), extrinsic incubation period in mosquitoes, blood meal digestion period, and finally mosquitoes- to-human transmission (2^nd^ bite) ^28^. Ideally, for DENV, the generation interval completes at around 16 days at 28-32 °C ^28^.

Bangladesh’s dengue season is characterized by hot and wet periods running between June to August. This is the period with the highest amount of rainfall in the country facilitating Aedes mosquito breeding in the country ^29^. The monthly mean growth factor above 1 for April – June indicates that for each of these months, the incidence of dengue cases will surpass the current month. Thus, we suggest starting vector control intervention in April in Bangladesh.

Globally and regionally in South and Southeast Asia, dengue cases are increasing. DENV infection increased by more than 46% between 2015 and 2019 in Region ^8^. In 2023, up until 31 May, a total of 1515,460 DENV infections were recorded in Brazil with 387 deaths^30^. In Malaysia, a total of 43,619 DENV infections have been recorded by 21 May 2023^30^. We found an increasing trend of DENV infection in Bangladesh. This increasing trend was much stiffer after the serotype DENV-3 was introduced in the country in 2018 ^12^. This increased trend is possibly linked with climate change in the region attributed to increased temperature and unusual rainfall, urbanization, population growth, inadequate water supply and storage practice, poor sewer, and waste management system, rise in global commerce and tourism ^8^.

The case fatality ratio (CFR) of primary dengue infection is very low with an estimation of 0.018% - 0.1% ^31^. However, the CFR of secondary dengue infection is high, although precise estimates are not available, some studies show more than 1% and reaching up to 4% ^32^. Bangladesh’s overall CFR of dengue infection (0.34%) seems slightly higher considering the overall CFR reported in other South and Southeast Asian countries ^8^. However, more than 69% of dengue-related deaths in Bangladesh were recorded after the introduction of the serotype DENV-3 in 2019. Thus, secondary infection is likely contributing to higher dengue-related deaths in Bangladesh. In addition, the CFR of the dengue virus infection might have been affected by a lack of active surveillance and missing the mild and asymptomatic cases, and not recording the cases outside the public hospital and few selected private hospitals in Bangladesh or weaker health care system in the country^12^. In some years, the CFR was very high, for example, in the year 2003, the CFR was 2.1 (total cases 486), in the year 2000, 1.68 (total cases 5,551), and in 2022, 0.45 (total cases 62,382). On the other hand, the CFR decreased in the second decade. This improvement is probably associated with improved access to the health care system, a better understanding of the treatment protocol including the availability of clinical management guidelines and training for the health care providers, better availability of Information, Education, and Communication (IEC) materials, community engagement and expansion of surveillance system to more hospitals in the surveillance system across the county in the recent years, and overall improvement of the economic condition of the country ^33–35^.

Two large dengue outbreaks occurred in Bangladesh in the year 2019 and 2022 both characterized by unusual weather patterns and the occurrence of two different serotypes. The 2019 outbreak was characterized by early rainfall of 120 mm in February compared to a monthly mean of 20 mm precipitation, along with the introduction of a new serotype of DENV-3 in the country ^12^. The 2022 outbreak was characterized by the late onset of rainfall with 297 mm rainfall in October compared to a monthly mean of 156 mm, and thus prolongation of vector transmission season along with the introduction of a new serotype, DENV-4 in the country ^29^. The occurrence of a new serotype exposed a large naïve population in a densely populated country like Bangladesh. A large proportion of the population is already infected with one of the serotypes of DENV with more than 80% of people living in Dhaka having antibodies against DENV ^24^. Another study predicted an estimated 40 million people being infected with DENV nationally and 2.4 million annual infections ^36^. Thus, any subsequent infections raise the risk of developing severe dengue hemorrhagic fever through antibody-dependent enhancement (ADE) ^7^. The deaths of many people in the year 2022 when the new serotype DENV-4 was introduced were probably associated with secondary dengue infection.

Controlling vector-borne diseases in tropical countries where temperatures, humidity, and rainfall remain favorable for breeding mosquitoes during most periods of the year is a difficult task^29^. Concerns were raised over the development of insecticide resistance ^12,37^ and the failure of developing a successful dengue vaccine ^38^. The prospect of *Wolbachia-*related intervention is bright but still far from applying on a national scale considering the expenses and technicalities associated with this. In this situation, an integrated and holistic vector management plan while engaging the local communities is key for controlling Aedes-borne diseases, especially in resource-limited countries. Regular destruction of mosquito breeding sites and increasing surveillance for detecting active cases are key in controlling dengue virus infection. Continuous active dengue surveillance will enable early detection of cases and outbreaks. Public health authorities will be able to identify areas where the disease is spreading, take immediate action to control mosquito populations, isolate infected patients, and implement public awareness campaigns to educate people about preventive measures. Early detection and response can help prevent the further spread of the disease and reduce its impact on individuals and communities.

Regular destruction of mosquito breeding habitats and increasing surveillance for detecting active cases should prioritize in controlling dengue virus infection in Bangladesh. Policymakers need to design an Aedes-borne disease management plan by considering a range of diseases that Aedes mosquito can transmit including Chikungunya, yellow fever, Zika virus, West Nile, Japanese Encephalitis, Eastern Equine Encephalitis, Ross River, Rift Valley fever, and the LaCrosse virus ^29^.

Our data should be viewed instead of several weaknesses of our study. We relied on the reported number of cases from the Ministry of Health and Family Welfare’s website, which mainly relies on passive reporting systems from the selected health facilities in the country^12^. These numbers seem to be an underestimation of actual cases. A modeling study based on the national seroprevalence of DENV antibodies predicted an annual infection of 2.4 million cases ^39^. However, dengue infection is underestimated globally as it is difficult to diagnose asymptomatic or mild cases that never reach healthcare settings. Although mild cases are missed more frequently, the severe and fatal cases would likely visit the hospital and thus be counted as numerators in our estimation. Thus, our estimation did not overlook the worst-case scenario, that is, our estimation, for example, estimated the higher CFR rather than the lower possible estimates.

## Conclusions

Between the first (2000-2010) and the second decade (2011-2022), DENV infection have increased by 8.3 times, and annual deaths have doubled in Bangladesh. This growth of DENV infection is partly explained by the influence of global warming with an increase of 0.49°C annual temperature as well as changes in duration and length of the rainy season. Unusual rain including early or late rain in and beyond the monsoon season likely contributed to extending the length of the dengue transmission season in Bangladesh. The monthly mean temperature, and monthly total rainfall of the first-and -second lagged months showed a greater influence on the monthly incidence of DENV infection in Bangladesh. The mean monthly growth factor remains significantly above one during April-July, which coincides with the hot and rainy season of the country indicating an earlier vector control would benefit the country. The ARIMA model forecasts a continuously increasing trend of DENV infection for the next decade in Bangladesh. We recommend an integrated and holistic vector management plan while engaging the local communities in the regular destruction of mosquito breeding sites and increasing surveillance for detecting active DENV-infected cases. Proactive surveillance, vector control, and vaccine rollout remain essential public health interventions. In the context of climate change, urbanization, trade, and the movement of people with vectors, there is a need to operationalize the One Health approach to address dengue fever and other vector-borne diseases in Bangladesh and beyond.

## Acknowledgments

We are grateful to the Ministry of Health and Family Welfare of Bangladesh for publicly sharing the dengue cases data. We acknowledge Bangladesh Meteorological Department for sharing the meteorological data. NH, and AZ, are part of the PANDORA-ID-NET Consortium (EDCTP 373 Reg/Grant RIA2016E-1609) funded by the European and Developing Countries Clinical Trials Partnership (EDCTP2) programme. NH is a member of the International Development Research Centre, Canada’s grant on West African One Health Actions for understanding, preventing, and mitigating outbreaks (109810-001). AZ is a National Institutes of Health Research senior investigator, and a Mahathir Science Award and Pascoal Mocumbi Award laureate.

## Author contribution statement

NH ideated the study and all authors helped develop the study outline and protocol. MNH and IK collected the data. NH, MNH, MA and AZ analyzed the data. NH, IK and MNH prepared the first draft manuscript and all authors contributed to several drafts and finalization of the manuscript. All authors approved the final draft and submission of the manuscript.

## Financial Support

There was no funding for this research.

## Conflict of interest

The authors declare that they have no conflict of interest.

## Ethics statement

This study does not include any individual-level data and thus does not require any ethical approval. We used publicly available data on Dengue cases and deaths.

## Data availability statement

All the dengue data presented in this manuscript are publicly available on Bangladesh’s Ministry of Health and Family Welfare’s Directorate General of Health Services website (https://dghs.gov.bd/). The meteorological data were purchased from Bangladesh Meteorological Department and are restricted to use for research purposes only and anyone interested in these data can request Bangladesh Meteorological Department (https://live3.bmd.gov.bd/).

## Notes

### Competing Interest Statement

The authors have declared no competing interest.

### Funding Statement

There was no funding for this study

## References

1 WHO. DENGUE: GUIDELINES FOR DIAGNOSIS, TREATMENT, PREVENTION AND CONTROL. Geneva, Switzerland, 2009.

2 CDC. Transmission through mosquito bites. 2019. https://www.cdc.gov/dengue/transmission/index.html (accessed May 15, 2023).

3 WHO. Dengue and severe dengue. https://www.who.int/news-room/fact-sheets/detail/dengue-and-severe-dengue. WHO. 2023; : 1–3.

4 WHO. Dengue and severe dengue. 2023. https://www.who.int/news-room/fact-sheets/detail/dengue-and-severe-dengue (accessed May 15, 2023).

5 Murray NEA, Quam MB, Wilder-Smith A. Epidemiology of dengue: past, present and future prospects. Clin Epidemiol 2013; 5: 299–309.

6 WHO-Bangladesh. Dengue - Bangladesh. https://www.who.int/emergencies/disease-outbreak-news/item/2022-DON424. 2022; published online Nov 28.

7 Teo A, Tan HD, Loy T, Chia PY, Chua CLL. Understanding antibody-dependent enhancement in dengue: Are afucosylated IgG1s a concern? PLoS Pathog 2023; 19: e1011223.

8 WHO South-East Asia. Dengue in the South-East Asia. WHO Regional office for South-East Asia. 2023; : 1–6.

9 Sharmin S, Viennet E, Glass K, Harley D. The emergence of dengue in Bangladesh: epidemiology, challenges and future disease risk. Trans R Soc Trop Med Hyg 2015; 109: 619–27.

10 Mutsuddy P, Tahmina Jhora S, Shamsuzzaman AKM, Kaisar SMG, Khan MNA, Dhiman S. Dengue Situation in Bangladesh: An Epidemiological Shift in terms of Morbidity and Mortality. Can J Infect Dis Med Microbiol 2019; 2019. DOI:10.1155/2019/3516284.

11 DGHS. DGHS. 2023. https://old.dghs.gov.bd/index.php/bd/home/5200-daily-dengue-status-report (accessed May 15, 2023).

12 Ahsan A, Haider N, Kock R, Benfield C. Possible Drivers of the 2019 Dengue Outbreak in Bangladesh: The Need for a Robust Community-Level Surveillance System. J Med Entomol 2021; 58: 37–9.

13 BMD. Climate and Weather Data Portal | Bangladesh Meteorological Department. 2023. http://www.bmddataportal.com/#/ (accessed May 15, 2023).

14 Haider N, Chang Y-M, Rahman M, Zumla A, Kock RA. Dengue outbreaks in Bangladesh: Historic epidemic patterns suggest earlier mosquito control intervention in the transmission season could reduce the monthly growth factor and extent of epidemics. Current Research in Parasitology & Vector-Borne Diseases 2021; 1: 100063.

15 Kumar N, Susan S. COVID-19 Pandemic Prediction using Time Series Forecasting Models. In: 2020 11th International Conference on Computing, Communication and Networking Technologies (ICCCNT). IEEE, 2020: 1–7.

16 Hasan MN, Haider N, Stigler FL, et al. The Global Case-Fatality Rate of COVID-19 Has Been Declining Since May 2020. Am J Trop Med Hyg 2021; 104: 2176–84.

17 Yue S, Pilon P. A comparison of the power of the t test, Mann-Kendall and bootstrap tests for trend detection / Une comparaison de la puissance des tests t de Student, de Mann-Kendall et du bootstrap pour la détection de tendance. Hydrological Sciences Journal 2004; 49: 21–37.

18 Sen PK. Estimates of the Regression Coefficient Based on Kendall’s Tau. J Am Stat Assoc 1968; 63: 1379–89.

19 Sumi SN, Sinha NC, Islam MA. Generalized linear models for analyzing count data of rainfall occurrences. SN Appl Sci 2021; 3: 481.

20 R Core Team. R: A language and environment for statistical computing. Vienna, Austria. URL https://www.R-project.org/. 2022.

21 Haider N, Rahman MS, Khan SU, et al. Identification and Epidemiology of a Rare HoBi-Like Pestivirus Strain in Bangladesh. Transbound Emerg Dis 2014; 61: 193–8.

22 Focks DA, Daniels E, Haile DG, Keesling JE. A simulation model of the epidemiology of urban dengue fever: Literature analysis, model development, preliminary validation, and samples of simulation results. American Journal of Tropical Medicine and Hygiene 1995. DOI:10.4269/ajtmh.1995.53.489.

23 Soneja S, Tsarouchi G, Lumbroso D, Tung DK. A Review of Dengue’s Historical and Future Health Risk from a Changing Climate. Curr Environ Health Rep 2021; 8: 245–65.

24 Salje H, Morales I, Gurley ES, Saha S. Seasonal Distribution and Climatic Correlates of Dengue Disease in Dhaka, Bangladesh. Am J Trop Med Hyg 2016; 94: 1359–61.

25 Rahman KM, Sharker Y, Rumi RA, et al. An Association between Rainy Days with Clinical Dengue Fever in Dhaka, Bangladesh: Findings from a Hospital Based Study. Int J Environ Res Public Health 2020; 17: 9506.

26 Cuong HQ, Hien NT, Duong TN, et al. Quantifying the Emergence of Dengue in Hanoi, Vietnam: 1998–2009. PLoS Negl Trop Dis 2011; 5: e1322.

27 Wangdi K, Clements ACA, Du T, Nery SV. Spatial and temporal patterns of dengue infections in Timor-Leste, 2005–2013. Parasit Vectors 2018; 11: 9.

28 Siraj AS, Oidtman RJ, Huber JH, et al. Temperature modulates dengue virus epidemic growth rates through its effects on reproduction numbers and generation intervals. PLoS Negl Trop Dis 2017. DOI:10.1371/journal.pntd.0005797.

29 Haider N, Hasan MN, Khalil I, et al. The 2022 dengue outbreak in Bangladesh: hypotheses for the late resurgence of cases and fatalities. J Med Entomol 2023; published online May 18. DOI:10.1093/jme/tjad057.

30 European CDC. Dengue worldwide overview. ECDC. 2023; : 1–10.

31 Huits R, Schwartz E. Fatal outcomes of imported dengue fever in adult travelers from non-endemic areas are associated with primary infections. J Travel Med 2021; 28. DOI:10.1093/jtm/taab020.

32 Liu Y, Lillepold K, Semenza JC, Tozan Y, Quam MBM, Rocklöv J. Reviewing estimates of the basic reproduction number for dengue, Zika and chikungunya across global climate zones. Environ Res. 2020. DOI:10.1016/j.envres.2020.109114.

33 Diseases Control Division (DGHS). National guidelines for clinical management of Dengue syndrome. Dhaka, 2013.

34 WHO. Improving the quality of care in the public health system in Bangladesh: building on new evidence and current policy levers: Bangladesh Health Systems in Transition, Policy Notes. Dhaka, 2017.

35 Albis MLF, Bhadra SK, Chin B. Impact evaluation of contracting primary health care services in urban Bangladesh. BMC Health Serv Res 2019; 19: 854.

36 Salje H, Paul KK, Paul R, et al. Nationally-representative serostudy of dengue in Bangladesh allows generalizable disease burden estimates. Elife 2019; 8. DOI:10.7554/ELIFE.42869.

37 Al-Amin HM, Johora FT, Irish SR, et al. Insecticide resistance status of Aedes aegypti in Bangladesh. Parasit Vectors 2020; 13: 622.

38 Wang TT, Sewatanon J, Memoli MJ, et al. IgG antibodies to dengue enhanced for FcγRIIIA binding determine disease severity. Science (1979) 2017; 355: 395–8.

39 Salje H, Paul KK, Paul R, et al. Nationally-representative serostudy of dengue in Bangladesh allows generalizable disease burden estimates. Elife 2019; 8. DOI:10.7554/eLife.42869.

